# Transmission potential of COVID-19 in Iran

**DOI:** 10.1101/2020.03.08.20030643

**Authors:** Kamalich Muniz-Rodriguez, Isaac Chun-Hai Fung, Shayesteh R. Ferdosi, Sylvia K. Ofori, Yiseul Lee, Amna Tariq, Gerardo Chowell

## Abstract

We estimated the reproduction number of 2020 Iranian COVID-19 epidemic using two different methods: R_0_ was estimated at 4.4 (95% CI, 3.9, 4.9) (generalized growth model) and 3.50 (1.28, 8.14) (epidemic doubling time) (February 19 - March 1) while the effective R was estimated at 1.55 (1.06, 2.57) (March 6-19).

## Main text

Iran has been experiencing a devastating epidemic of COVID-19 in 2020 (1). The virus appears to be spreading rapidly with multiple cases reported among the elite (2). One model estimated that 18,300 (95% CI, 3,770 to 53,470) cases would have had occurred by February 24 (3). The Iranian authorities have adopted social distancing measures to slow disease transmission (4). As the epidemic continues, our results call for efforts to minimize COVID-19-related morbidity and mortality.

To confront the epidemic, the transmission potential of COVID-19 is a useful metric to guide the outbreak response efforts including the scope and intensity of interventions. During the early transmission phase, the basic reproduction number, R_0_, represents such a measurement quantifying the average number of secondary cases that primary cases generate in a completely susceptible population in the absence of interventions or behavioral changes. R_0_>1 indicates the possibility of sustained transmission, whereas R_0_<1 implies that transmission chains cannot sustain epidemic growth. As the epidemic continues its course, the effective reproduction number, R_e_, offers a time-dependent record of the average number of secondary cases per case as the number of susceptible individuals gets depleted and control interventions takes effect. Herein we employed two different methods to quantify the reproduction number using the curve of reported COVID-19 cases in Iran and its five regions (Table S1).

Using a Wikipedia entry as a starting point, Farsi-speaking coauthor SFR double-checked the data of daily reported new cases, from February 19 through March 19, 2020 (the day before the Iranian New Year), using Iranian official press releases and other credible news sources, and corrected the data as per the official data (Figure S1, Tables S2 and S3). Incident cases for the 5 regions were missing for two days: March 2-3, 2020, which were not included in our analysis. We noted that the reported national number of new cases did not match the sum of the new cases reported in Iran’s five regions on March 5, 2020; we decided to treat each time series as independent and used the data as reported. Using Method 1, we estimated R_0_ using data from February 19 through March 1, 2020. Using Method 2, we estimated R_0_ from the early transmission phase (February 19 through March 1, 2020) and R_e_ based on the growth rate estimated from March 6 through 19, 2020 during which the epidemic slowed down likely reflecting the impact of social distancing measures.

Two methods were utilized to estimate the reproduction number. The first method utilized a generalized growth model (GGM) (5) with the growth rate and its scaling factor to characterize the daily reported incidence, followed by simulation of the calibrated GGM, using a discretized probability distribution of the serial interval and assuming a Poisson error structure. See Technical Appendix for detailed description of Method 1.

The second method for estimating the reproduction number is based on the calculation of the epidemic’s doubling times, which correspond to the times when the cumulative incidence doubles. We estimated the epidemic doubling time using the curve of cumulative daily reported cases. To quantify parameter uncertainty, we used parametric bootstrapping with a Poisson error structure around the number of new reported cases in order to derive 95% confidence intervals of our estimates (6-8).

Assuming exponential growth, the epidemic growth rate is equal to ln(2)/doubling time. Assuming that the pre-infectious and infectious periods follow an exponential distribution, R_0_ is approximately equal to 1 + growth rate × serial interval. See Technical Appendix and Equation 4.14, Table 4.1 in Vynnycky and White (9).

In both methods, the serial interval was assumed to follow a gamma distribution (mean: 4.41 days; standard deviation: 3.17 days) (10, 11). MATLAB version R2019b and R version 3.6.2 were used for data analyses and creating figures. It was determined *a priori* that α= 0.05.

**Figure.**
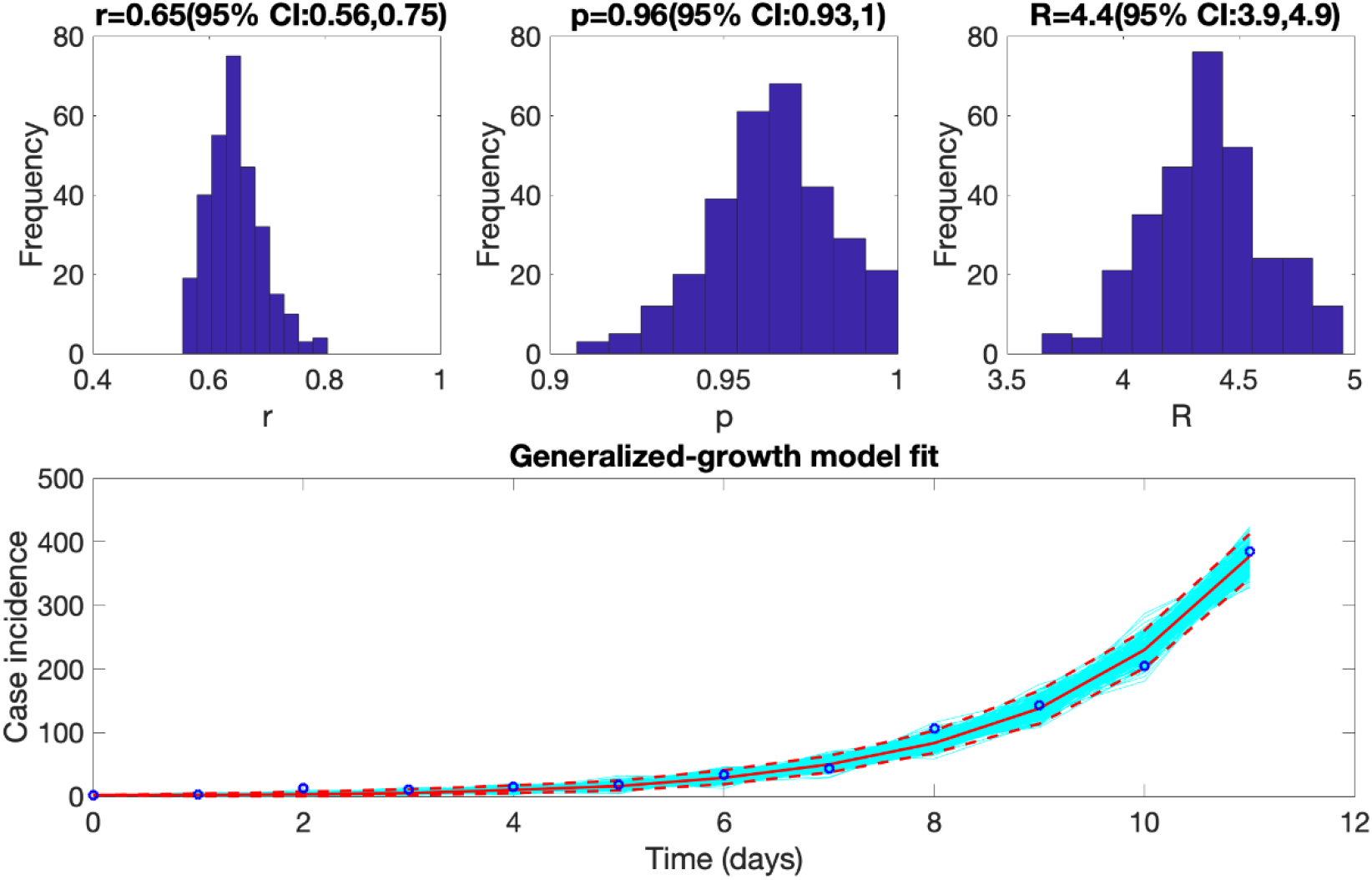
The mean basic reproduction number of COVID-19 epidemic in Iran, with 95% confidence interval. Estimates for growth rate, r, and the scaling of the growth rate parameter, p, are also provided. The plot in the lower panel depicts the fit of the Generalized Growth Model (Method 1) to the Iranian data assuming Poisson error structure as of March 1, 2020.

Using Method 1, we estimated an R_0_ of 4.4 (3.9, 4.9) for the COVID-19 epidemic in Iran. We estimated a growth rate of 0.65 (95% CI, 0.56, 0.75) and a scaling parameter of 0.96 (95% CI, 0.93, 1) (Table S4). The latter indicated a near-exponential growth of the epidemic (**Figure**). Using Method 2, we found that from February 19 through March 1, the cumulative incidence of confirmed cases in Iran had doubled 8 times. The estimated epidemic doubling time was 1.20 (95% CI, 1.05, 1.45) days and the corresponding R_0_ estimate was 3.50 (95% CI, 1.28, 8.14). From March 6 through March 19, the cumulative incidence of confirmed cases had doubled 1 time, with a doubling time of 5.46 (5.29, 5.65) days. The corresponding effective R estimate was 1.55 (95% CI, 1.06, 2.57) (Table S5, Figures S7, S8). Our results are robust, as they are consistent with the Iranian COVID-19 R_0_ estimates of 4.7 and 4.86 generated by Ahmadi et al. and Sahafizadeh and Sartoli (12, 13), respectively, but are higher than the R_0_ of 2.72 estimated by Ghaffarzadegan & Rahmandad (14).

Our study has limitations. First, our analysis is based on the number of daily reported cases whereas it would be ideal to analyze case counts by date of symptoms onset, which were not available. Second, case counts could be underreported due to underdiagnosis, given subclinical or asymptomatic cases, or limited testing capacity to test mild cases. The rapid increase in case counts might represent a belated realization of the severity of the epidemic and a rapid process of catching up with testing many suspected cases. If the reporting ratio remains constant over the study period, and given the near-exponential growth of the epidemic’s trajectory, our estimates would remain reliable; however, this is a strong assumption. Third, while data are not stratified according to imported and local cases, we assumed that they were infected locally, as it is likely that transmission has been ongoing in Iran for some time (3).

In conclusion, we used two different methods to compute the R_0_ of COVID-19 epidemic in Iran. Our mean estimate was at 4.4 (95% CI, 3.9, 4.9) for Method 1, and at 3.50 (95% CI, 1.28, 8.14) for Method 2, which means that on average around four susceptible individuals were infected by one infectious individual during the early growth phase. When social distancing interventions are in place, the epidemic slows. The doubling time of the COVID-19 epidemic in Iran increased from 1.20 (95% CI, 1.05, 1.45) to 5.46 (95% CI, 5.29, 5.65) days, and the effective R dropped to 1.55 (95% CI, 1.06, 2.57). While the COVID-19 epidemic in Iran has now slowed down substantially, the situation in Iran is still dire with the average daily number of ∼2290 new reported cases over 14 days from March 20 through April 2, 2020. Tightening social distancing interventions is needed to bring the epidemic under control in Iran.

## Data Availability

The data used in this paper is publicly available. We also provide the data in the Technical Appendix.

## Co-first authors’ biographies

Kamalich Muniz-Rodriguez, a doctoral student in epidemiology, and Dr. Isaac Chun-Hai Fung, an associate professor of epidemiology, are at Jiann-Ping Hsu College of Public Health, Georgia Southern University. Their research interests include infectious disease epidemiology, digital health and disaster emergency responses.

## Acknowledgement

GC acknowledges support from NSF grant 1414374 as part of the joint NSF-NIH-USDA Ecology and Evolution of Infectious Diseases program. ICHF acknowledges salary support from the National Center for Emerging and Zoonotic Infectious Diseases, Centers for Disease Control and Prevention (19IPA1908208). This article is not part of ICHF’s CDC-sponsored projects.

## Disclaimer

This article does not represent the official positions of the Centers for Disease Control and Prevention, the National Institutes of Health, or the United States Government.

## Ethics statement

The Georgia Southern University Institutional Review Board has made a non-human subjects determination for our project (H20364), under G8 exemption category.

